# Extended interval BNT162b2 vaccination enhances peak antibody generation in older people

**DOI:** 10.1101/2021.05.15.21257017

**Authors:** H Parry, R Bruton, C Stephens, K Brown, G Amirthalingam, B Hallis, A Otter, J Zuo, P Moss

**Affiliations:** Institute of Immunology and Immunotherapy, University of Birmingham, Birmingham. B15 2TT, UK; National infection Service, Public Health England, Colindale, London NW9 5EQ, UK; National infection Service, Public Health England, Porton Down, Salisbury, SP4 OJG, UK

**Keywords:** Vaccination, SARS-CoV-2, COVID, Immunosenescence, Antibody, T-cell, Ageing

## Abstract

**Objectives:** To assess the relative immunogenicity of standard or extended interval BNT162b2 vaccination.

**Design:** Population based cohort study comparing immune responses 2 weeks after the second vaccine, with appropriate time-matched samples in participants who received standard or extended interval double vaccination.

**Setting:** Primary care networks, Birmingham, UK. December 2020 to April 2021.

**Participants:** 172 people aged over 80 years of age. All donors received the BNT162b2 Pfizer/BioNTech vaccination and were vaccinated with either a standard 3 week interval between doses or an extended interval schedule.

**Main outcome measures:** Peak quantitative spike-specific antibody and cellular immune responses.

**Results:** In donors without evidence of previous infection the peak antibody response was 3.5-fold higher in donors who had undergone delayed interval vaccination. Cellular immune responses were 3.6-fold lower.

**Conclusion:** Peak antibody responses after the second BNT162b2 vaccine are markedly enhanced in older people when this is delayed to 12 weeks although cellular responses are lower. Extended interval vaccination may therefore offer the potential to enhance and extend humoral immunity. Further follow up is now required to assess long term immunity and clinical protection.

**What is already known on this topic:** The BNT162b2 vaccine is highly effective against Covid-19 infection and was delivered with a 3-week time interval in registration studies. However, this interval has been extended in many countries in order to extend population coverage with a single vaccine. It is not known how immune responses after the second dose are influenced by delaying the second vaccine.

**What this study adds:** We provide the first assessment of immune responses in the first 14 weeks after standard or extended interval BNT162b2 vaccination and show that delaying the second dose acts to strongly boost the peak antibody response in older people. The extended interval vaccination may offer a longer period of clinical protection. This information will be of value in optimizing vaccine regimens and help guide guide vaccination policies.

## Introduction

SARS-CoV-2 vaccines have demonstrated remarkable efficacy in protection against infection and symptomatic disease(1). As such they offer the potential to provide large scale protection against the Covid-19 pandemic but many questions remain regarding their optimal delivery for provision of effective and sustained immunity. The clinical severity of SARS-CoV-2 infection is increased in older people and as such this group has been prioritised for vaccination in most countries(2). However, the quality of immune responses to vaccination deteriorates with age due to immunosenescence and as such there is considerable interest in understanding how to optimise vaccine schedules within this age group in order to maximise protection (3).

The BNT162b2 mRNA vaccine is authorised for administration with a three week interval between the two doses. However several countries have chosen to delay the timing of the second vaccine by up to 12 weeks in order to accelerate the proportion of people that have received at least one vaccine dose and enhance population protection. Real world evidence now indicates that this strategy is highly effective(4). Our previous work has shown that over 90% of people over the age of 80 years develop antibody responses at 5 weeks following a single BNT162b2 vaccine(5). However, it is not yet known how well antibody and cellular responses are boosted after the second vaccine in patients of this age group on extended-interval vaccine regimens.

Here we compared spike-specific antibody and cellular immune responses in a large population of older people who underwent dual BNT162b2 vaccination with either a 3-week or 11-12 interval. We demonstrate that both approaches generate high levels of antibody response but peak values are 3.5-fold higher with the extended-interval protocol. In contrast, cellular specific responses were stronger in people who had the standard 3-week vaccine interval. As such the timing of the second vaccine has a marked influence on the kinetics and magnitude of the adaptive immune response after mRNA vaccination in older people. These considerations should be of value in optimisiming strategies that provide long term protection against SARS-CoV-2 infection.

## Methods

### Participants

172 participants aged 80 years and older, and who were living independently, were recruited to study. The work was performed under the CIA UPH IRAS approval (REC 20\NW\0240) and conducted according to the Declaration of Helsinki and good clinical practice.

All donors received the BNT162b2 Pfizer/BioNTech vaccination and were vaccinated with a standard 3 week interval between doses (median age 84 years (IQR 80-87 or range 80-96) or an extended interval schedule, with the second vaccine given 11-12 weeks after the first (median age 84 years, IQR 82-89, range 80-99). Participants received the same phlebotomy time points at 5-6 weeks and 13-14 weeks following the first vaccine, for comparative purposes.

### Roche Elecsys® electrochemiluminescence immunoassay (ECLIA)

Serum was stored at -20^0^C and defrosted prior to antibody analysis. IgG/A/M antibodies specific to SARS-CoV-2 were detected using electrochemiluminescence assays on the automated Roche cobas e801 analysers based at Public Health England (PHE) Porton. Calibration and quality control were performed as recommended by the manufacturer. Anti-nucleocapsid protein (NP) antibodies were detected using the qualitative Roche Elecsys® AntiSARS-CoV-2 ECLIA (COV2, Product code: 09203079190), whilst anti-spike (S) antibodies were detected using the quantitative Roche Elecsys® Anti-SARS-CoV-2 S ECLIA (COV2 S, Product code 09289275190). Anti-nucleocapsid results are expressed as cut-off index (COI) value, with a COI value of ≥1.0 considered positive for anti-nucleocapsid antibodies. Anti-spike results are expressed as units per ml (U/ml), with samples with a result of ≥0.8 U/ml considered positive for anti-spike antibodies within the fully quantitative range of the assay: 0.4 – 2,500 U/ml. Samples >2,500 U/ml were diluted further (1:10, 1:100 and 1:1000) to within the quantitative range.

### Cellular assays

Peripheral blood mononuclear cells (PBMCs) were isolated from a whole blood sample using ‘T-Cell Xtend’ (Oxford Immunotec) and Ficoll. After quantification and dilution of recovered cells, 250,000 PBMC were plated into each well of a ‘T-SPOT Discovery SARS-CoV-2’ kit (Oxford Immunotec). This is designed to measure responses to overlapping peptides pools covering protein sequences of four different SARS-CoV-2 antigens, without HLA restriction, and includes negative and positive controls. Peptide sequences that showed high homology to endemic coronaviruses were removed from the sequences, but sequences that may have homology to SARS-CoV-1 were retained. Cells were incubated and interferon-γ secreting T cells were counted. A cut off of 6+ spots per 250,000 PBMCs on the S1 pool was defined as a positive response in line with the Oxford Immunotec diagnostic Covid kit.

### Statistical analysis

Data were tested for normality using Kolmogorov-Smirnov analysis. For comparative analysis of antibody titres and cellular responses within the same cohort Wilcoxon ranked pairs was performed. For comparative analysis of antibody or cellular responses between the 2 cohorts, Mann-Whitney t-test was performed. Spearman’s rank correlation was used to assess the relationship with cellular response and the rate of antibody waning. All analysis was performed using Graphpad prism v9.1.0 for Mac (San Diego, California USA).

## Results

### Standard-interval and extended-interval vaccination regimens elicit strong antibody responses but peak values are 3.5-fold higher with the extended-interval regimen

Analysis was undertaken in donors aged over 80 years who received two doses of the Pfizer/BioNTech BNT162b2 vaccine. In 99 participants the two doses were given at 3 weeks apart which we term the ‘standard interval’. In 73 participants the two doses were given at 11-12 weeks apart which we term the ‘extended interval’.

Venepuncture blood samples were taken at two timepoints in the standard and extended interval vaccine cohorts respectively. Within the standard interval cohort the first sample was taken at 2-3 weeks following the second vaccine to determine the ‘peak response’ to the vaccine boost. A second sample was taken 8-9 weeks after the second vaccine in order to assess the stability of antibody and cellular responses over this period (n=79).

For donors with an extended interval vaccine schedule, blood samples were taken at 5-6 weeks following the first dose vaccine (n=68) and then again at 8-9 weeks later. This second timepoint was 2-3 weeks after the second vaccine dose and therefore represented the ‘peak response’ in the extended interval cohort (n= 55) (figure 1).

**Figure 1.**
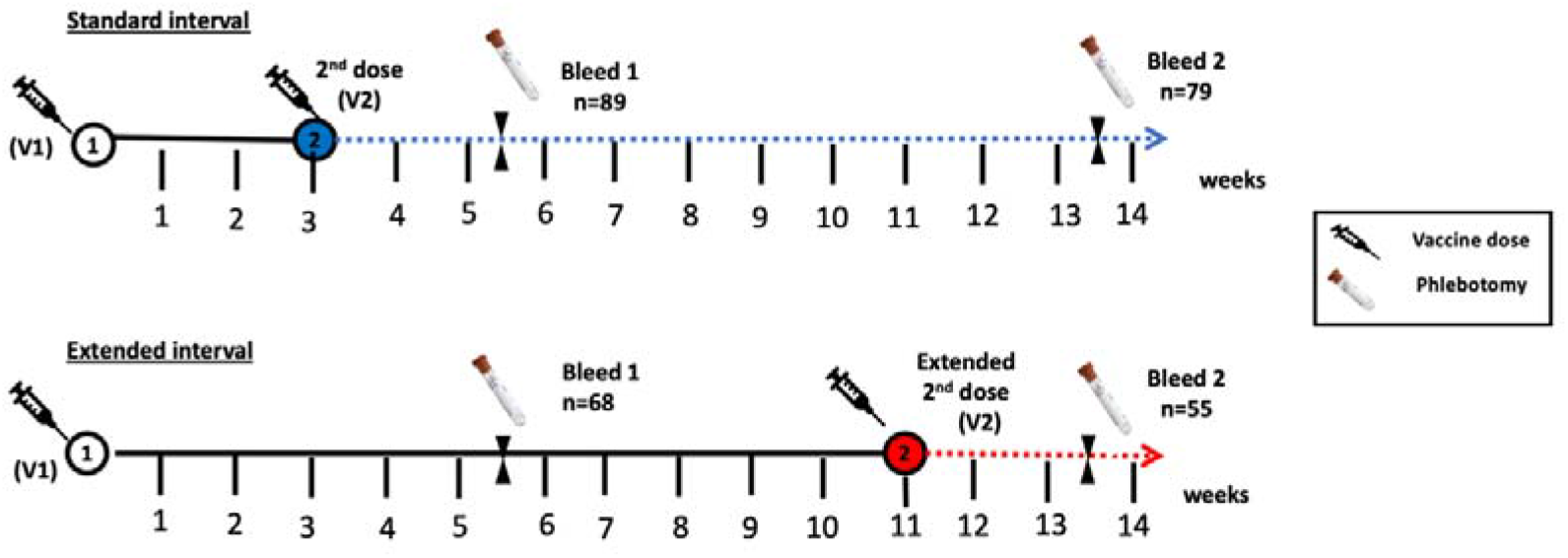
Infographic of vaccination protocol and blood collection timepoints. Blood samples were obtained on all participants at two timepoints. These were at 5-6 weeks and then again at 13-14 weeks. Bleed 1 corresponded to 2-3 weeks post second vaccine (V2) for the standard-interval cohort and 5-6 weeks post first vaccine (V1) in the extended-interval cohort. Bleed 2 corresponded to 10-11 weeks post second vaccine (V2) in the standard interval cohort and 2-3 weeks post second vaccine (V2) in the extended interval cohort. The number of study participants is shown and includes only those with no previous natural infection with SARS-CoV-2.

Initial studies were undertaken to determine the status of participants in relation to previous natural infection with SARS-CoV-2 through detection of nucelocapsid-specific antibodies. These were present in 10 and 5 donors in the standard and extended interval vaccine regimens respectively. As previous infection has a major impact on the immune response to vaccination these donors were excluded from primary analysis.

Spike-specific antibodies were detected in 100% of participants in the standard interval cohort at both the first and second timepoints (n=86 and n=79 respectively). Within the extended interval cohort, antibodies were detectable in 91% (62/68) at the first timepoint, at 5-6 weeks after the first vaccine, but this rose to 100% 2-3 weeks after the vaccine boost.

We next went on to assess the magnitude of the antibody response at the two timepoints within both cohorts. Antibody titres in the standard-interval regimen peaked at 1138 U/ml after the second dose and then fell by 2.6-fold over the subsequent weeks (p<0.0001) (figure 2A). Within the extended-interval cohort the median antibody titre was 17 U/ml at 5-6 weeks weeks after the first vaccine but showed a substantial 242-fold increase to reach 4030 after the second boost (p<0.0001) (figure 2B). The kinetics of antibody responses in the standard- and extended-interval cohorts are shown graphically in figure 2C.

**Figure 2.**
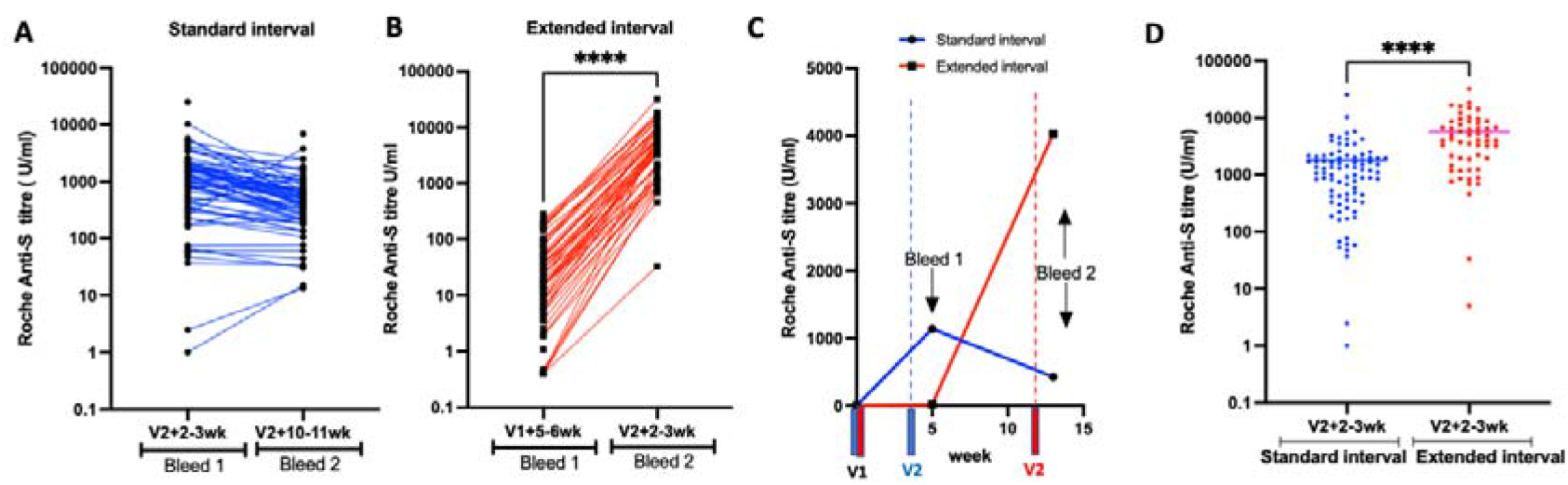
Extended-interval vaccination with BNT162b2 stimulates stronger peak spike-specific antibody responses. **(A)** Dot plot to compare spike-specific antibody responses in participants who received the BNT162b2 vaccine with a standard interval of 3 weeks, apart at time point 1(2-3 weeks post vaccine 2) and time point 2 (10-11 weeks post vaccine 2). **(B)** Dot plot to compare spike-specific antibody responses in participants who received the BNT162b2 vaccine with an extended interval at time point 1 (5-6 weeks post vaccine dose 1) and time point 2 (2-3 weeks post vaccine dose 2). **(C)** Kinetics of spike-specific antibody response during the 2 different vaccine interval schedules over a 14 week period. Blue lines represent participants in the standard 3 week interval and red lines represent those in the extended schedule. **(D)** Dot plot to compare spike-specific antibody responses at 2-3 weeks after the second dose BNT162b2 vaccine in the standard and extended interval cohorts.

When peak antibody responses after the second vaccine were contrasted in both cohorts it was apparent that values in the extended-interval group were 3.5-fold higher at 4030 U/ml compared to 1138 U/ml (p=<0.0001; Figure 2D).

### Peak cellular responses are higher in donors within the standard-interval vaccine regimen

Interferon-gamma (IFN-γ) ELISPOT analysis was then used to determine spike-specific T cell responses following vaccination in the two cohorts. Cellular responses against two peptide pools from the S1 and S2 spike domains were determined following overnight stimulation. Values from both wells were aggregated to give the total spike-specific response.

Within the standard-interval cohort 60% (53/89) of donors had a confirmed cellular response at 2-3 weeks following the second vaccine although this fell to only 15% (12/79) 8-9 weeks later. The proportion of participants demonstrating a cellular response in the extended-interval cohort was only 8% (5/67) at 5-6 weeks after the first vaccine but this rose to 31% (17/55) 2-3 weeks after the second vaccine (figure 3C).

**Figure 3.**
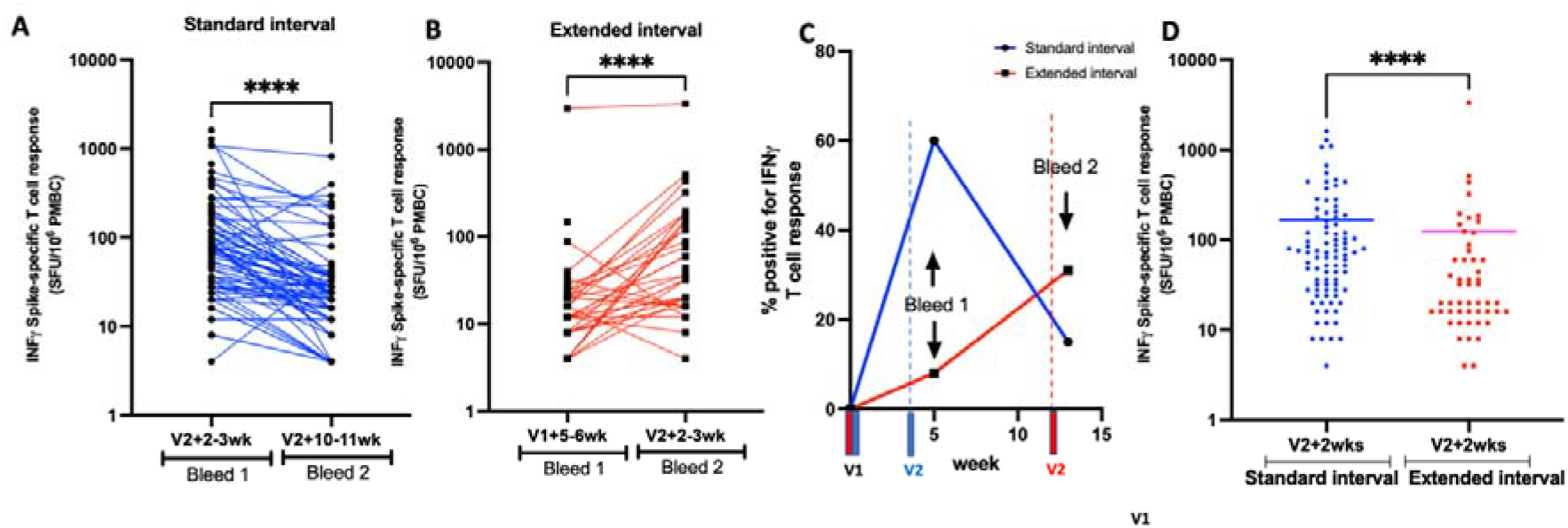
Standard interval vaccination with BNT162b stimulates a greater peak cellular response. **(A)** Dot plot to compare Spike-specific cellular responses by IFN-γ ELISpot in a participants who obtained the BNT162b2 vaccine with a standard interval of 3 weeks apart at bleed time point 1 (2-3 wks post vaccine dose 2) and time point 2 (10-11 wks post vaccine dose 2). **(B)** Dot plot to compare Spike-specific antibody responses by IFN-γ ELISpot in a participants who obtained the BNT162b2 vaccine with an extended interval at bleed time point 1 (5-6 weeks post vaccine dose 1) and time point 2 (2-3 weeks post vaccine dose 2). **(C)** The kinetics of the cellular response of the 2 different vaccine schedules are shown over a 14 week period. The blue lines represent those participants who obtained the BNT162b2 vaccine with a standard interval 3 weeks apart and the red lines represent those who received am extended interval to second dose vaccination of BNT162b2. **(D)** Dot plot to compare Spike-specific cellular responses by ELISpot in the participants 2-3 weeks after the second dose of BNT162b2 vaccination in the standard and extended interval cohorts.

The magnitude of spike-specific T cell responses in the standard-interval cohort peaked at 72 spots/million PBMC at 2-3 weeks following the second vaccine and then fell by 3-fold to 24 spots/million after 8-9 weeks (p<0.0001) (figure 3A). Within the extended-interval cohort these corresponding values were 8 spots/million 5-6 weeks after the first vaccine, followed by a 2.5-fold rise to reach 20 spots/million 2-3 weeks after the second vaccine (p<0.0001) (figure 3B).

A comparison of the median magnitude of peak cellular responses after the second vaccine in the two schedules showed that these were higher for donors in the standard-interval regime (72 vs 20 spots/million; p<0.0001). (figure 3D).

Cellular immune responses play an important role in supporting and maintaining antibody production and we therefore assessed the relationship between the cellular response 2 weeks following the second vaccine and the degree of subsequent antibody waning over the next 8-9 weeks in the standard-interval cohort. No association was found between S1 and S2-specific cellular responses and the rate of decline in antibody titre (r=-0.01; p=0.93).

## Discussion

Extended interval vaccination has been adopted in several countries and shown good clinical efficacy(4). Here we show that this approach leads to an enhanced peak antibody response after the second BNT162b2 vaccine although peak cellular responses were lower. These observations raise a number of questions regarding the underlying mechanisms of adaptive immunity to vaccination with potential relevance to vaccine strategy.

Spike-specific antibody responses within the first 2-4 weeks after second vaccine are emerging as a potential immune correlate of protection after vaccination(6). As such the strong humoral responses that are elicited by mRNA vaccines are likely to underlie their excellent clinical efficacy to date(7). Our findings confirm previous studies showing that the 3-week standard-interval BNT162b2 regimen elicits strong antibody responses in older people(8). As previously reported, we detected antibody responses in all donors at 2 weeks after the second vaccine. Here we were able to extend this work to assess the stability of adaptive immunity over the next two months. A median 2.6-fold reduction in antibody titre was seen over this 8 week period and indicates waning in the early post-boost period. However, systemic antibody levels would be expected to subside within the first few weeks following antigen challenge and absolute values remained substantial in most people(9). It will be important to assess how antibody levels are maintained over longer periods and this is likely to define the potential need for booster vaccines in this vulnerable age group.

There is no information to date regarding the impact of extended interval mRNA vaccination on peak antibody responses after the second dose in older people. Interestingly, we found that this approach boosted median peak antibody titres by 3.5-fold compared to those seen after standard regimen. Very high antibody levels were seen in a substantial proportion of donors with remarkable titres up to 18,100 U/ml. These responses are encouraging for long term protection and, although expected to reduce substantially over the subsequent few weeks, it is possible that this higher baseline may act to provide more robust long term protection. This may be of particular importance in relation to protection from viral variants of concern which may emerge as a major challenge for Covid-19 vaccinees(10). It will be of interest to assess the relative induction of long lived plasma cells and memory B cells following each regimen in order to assess potential cellular correlates of antibody response(11).

Prior natural infection with SARS-CoV-2 strongly enhances vaccine responses and median peak antibody responses of 32,250 U/ml were seen in the 10 donors who received the standard interval vaccination protocol. Of note, these values fell by 3.5-fold over the subsequent 8 weeks which is a greater rate of decline than in the infection-naive cohort (Supplementary Figure 1). This requires further longitudinal follow up in order to see if antibody levels plateau at values higher than in infection-naive donors. Of the 5 donors in the extended interval protocol who had prior natural infection, 2 had blood sampling at the second bleed point, with a peak antibody responses of 90,750 U/ml.

The importance of cellular immunity in providing clinical protection against SARS-CoV-2 is not currently clear. Our findings suggest that earlier administration of the second vaccine provides a greater boost to the cellular immune response. It is not clear why antibody and cellular responses show a differential response to standard and extended interval vaccination. mRNA vaccines lead to germinal centre formation and particularly strong induction of antibody responses(11, 12), but somewhat less is known regarding the induction of cellular immunity(13).

In both cohorts the proportion and magnitude of T cell response was somewhat lower than reported in younger cohorts and may reflect the impact of immunosenescence(14). It should be noted that T cell responses were measured on the standard assessment of IFN-γ secretion but this does not preclude the presence of spike-specific T cells which make other inflammatory cytokines and may be induced preferentially following mRNA vaccination(15). In particular, induction of T follicular helper cells often correlates with antibody induction. Indeed, one role for cellular immunity following SARS-CoV-2 vaccination may be to support the generation and maintenance of antibody but we did not find any evidence to suggest that the magnitude of cellular response following the second vaccine was associated with the rate of waning of antibody responses in the standard interval cohort.

Recent studies indicate that antibody levels remain robust for 6 months following 3-week double mRNA vaccination and decline with a half-life of 52 days after day 43(16). This is likely to underlie the impressive extended clinical efficacy over this time period(17). Our findings raise the question of whether or not the clinical efficacy of dual mRNA vaccination might be further enhanced by extending the interval between doses. It is noteworthy that an extended interval protocol for the adenovirus-based ChAdOx1 vaccine has also been shown to increase spike-specific antibody responses by 2.3-fold and to improve vaccine efficacy (18). The potential disadvantage of this approach is that it extends the period of partial protection prior to the second dose. However, epidemiological data indicates that single vaccination delivers strong clinical protection against symptomatic Covid-19 infection and as such this may not represent a major concern(4, 19). As such, if extended vaccine schedules act to establish a higher ‘baseline’ level of SARS-CoV-2-specific antibody then this may be worthy of consideration in relation to potentially minimizing the need for subsequent revaccination.

In conclusion we show that extended interval vaccination with BNT162b2 increases the peak antibody response by 3.5-fold in older people. This may help to sustain humoral immunity over the longer term and further improve the clinical efficacy of this powerful vaccine platform.

## Data Availability

Authors agree to share the anonymised raw data for this study once published.

## Author contributions

PM, HP wrote the manuscript. HP, RB, PM designed the study and HP, RB, KB, GA recruited participants. JZ, CS, AO, BH performed the experiments. HP, JZ analysed the data. All authors commented on the manuscript.

## Ethics approval

The work was performed under the CIA UPH IRAS approval and conducted according to the Declaration of Helsinki and good clinical practice. Ethical approval was obtained from North West Preston Research Ethics Committee with favourable outcome and approval (REC 20\NW\0240).

## Ethical Approval

The work was performed under the CIA UPH IRAS approval (REC 20\NW\0240) and conducted according to the Declaration of Helsinki and good clinical practice.

## Acknowledgements

We are very grateful to Dr Philip Saunders and Dr Rory Meade and the support from patients and staff at Lordswood Medical Group, Ridgacre House Surgery and Harborne Medical Practice. We also thank Millie Manning, Danielle Sutherland, Tamsin Drury and Alex Bray for their help in recruitment, as well as Rajinder Jeet and Shahada Rahman Joli with phlebotomy services.

---

**Supplementary:**
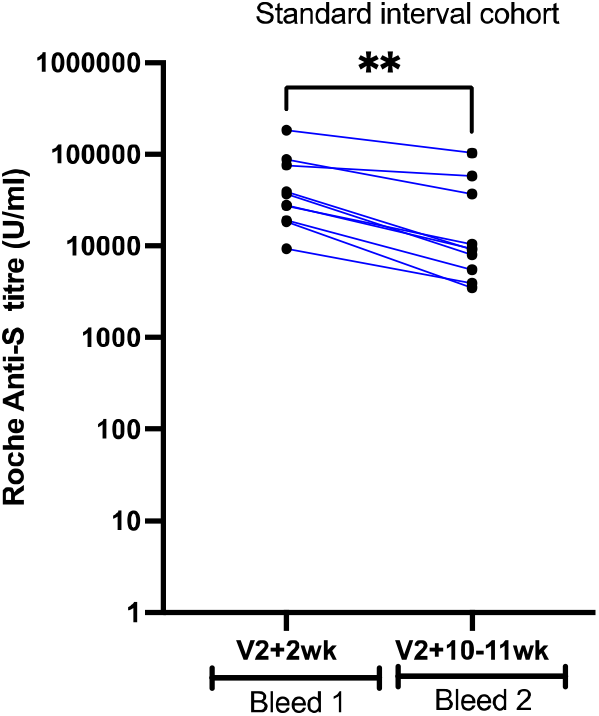
Dot plot to compare Spike-specific antibody responses by Roche platform in participants who were previously naturally exposed to SARS-CoV-2 and had obtained the BNT162b2 vaccine with a standard interval of 3 weeks apart (bleed time point 1 median 32250 U/ml: vs 9235 U/ml at bleed time point 2 median: p=0.002)

